# Exploring community-based reporting of livestock abortions for Rift Valley Fever surveillance in Uganda: A pilot study

**DOI:** 10.1101/2024.05.24.24307858

**Authors:** Abel Wilson Walekhwa, Andrew JK Conlan, Stella Acaye Atim, Anna Rose Ademun, Emmanuel Hasahya, James L.N. Wood, Lawrence Mugisha

## Abstract

Between March and June 2023, we carried out a pilot study to explore the feasibility of using self-reporting of livestock abortions as a method of syndromic surveillance for Rift Valley fever disease (RVF) in Isingiro District, Uganda. We established a call centre in the office of the District Veterinary Officer which was promoted through stakeholder meetings, media engagement and distribution of sensitization materials.

We collected 200 sera and 269 vaginal swabs from cattle, sheep and goats that experienced an abortion within a target 14-day period. The apparent IgG seroprevalence of RVF was 38% [95% CI 29 – 47] in cattle, 33% [95% CI 14 – 61] in sheep and 20% [95% CI 12 – 31] in goats. For IgM, sheep showed the highest prevalence at 8% [95% CI 1 – 35], cattle at 2% [95% CI 1 – 6]. Host species was significantly associated with IgG positive status, with cattle having the highest risk of RVF seropositivity (OR = 3 [95%CI: 1 - 7], p = 0.014).

Our results demonstrate the potential for a community led model for collecting abortion alerts through local call centres. If routinely implemented, such syndromic data collection could be used to develop early warning systems and prioritise case investigations. The IgG seroprevalence in our sample is comparable to the levels seen in endemically infected countries, suggesting historical circulation of RVF within the livestock population in this community. Our pilot study demonstrates a proof-of-principle that community-driven reporting of abortions could be used to build a surveillance system for RVF. However, the lack of confirmation of infection through PCR means we cannot draw a firm causal link between the reported abortions and RVF. To build a robust case of abortion surveillance, longitudinal studies are needed to measure seasonal variation in the distribution of abortion cases and incidence of active RVF infections.

**Author’s Summary:** Rift Valley fever disease is a growing zoonotic disease with high potential to disrupt national and international trade and biosecurity. In this work, we contribute to the epidemiology of RVF in Uganda and propose a system for self-reporting of livestock abortions that could provide a pathway to syndromic surveillance for RVF and thus contribute to safer human and animal communities. Engaging communities in setting up call centres for early notification and follow-up of animal diseases is a cost-effective and participatory approach which could be sustainable even with limited resources. Livestock abortions are a common symptom for other diseases like Q-fever, brucellosis, campylobacter among others and therefore, caution should be used when using it as an early warning signal for RVF. Establishing routine reporting could potentially allow for the identification and association of patterns of abortions with different infections. Our pilot study demonstrates the feasibility and active interest and engagement from farmers for such a surveillance system.

## Background

Rift valley fever disease (RVF) is a zoonotic mosquito-borne viral infection caused by the Rift valley fever virus (RVFV), a member of the genus *Phlebovirus* and family *Bunyaviridae* (Sasaya et al., 2023). The disease is primarily transmitted by *Aedes* and *culex* mosquitoes and blood feeding flies (Ebogo-Belobo et al., 2023). The virus was identified in 1931 during an epidemic among sheep on a farm in the Kenyan Rift Valley (Bailey, 1988; Hartman, 2017) affecting both livestock and wildlife (Clark et al., 2018). In humans, infection with RVFV leads to clinical outcomes ranging from a mild flu-like illness to severe haemorrhagic fevers with mortality rates of up to 27% (Ebogo-Belobo et al., 2023). The disease has the potential to disrupt countries’ livestock production due to mortalities (Ahmed et al., 2019). In leu of the potential for RVF to cause economic disruptions, the World Health Organisation (WHO) and Food Agricultural Organisation and World Organization for Animal Health (WOAH) have prioritized it among the key zoonotic diseases that have pandemic potential (Jenkin et al., 2023). The African Union Commission (AUC) has ranked it among the priority diseases for Africa, while in Uganda it is ranked third on the list of priority zoonotic diseases (PZDs) together with other viral haemorrhagic fevers (VHFs) including Ebola, Marburg and Crimean-Congo Haemorhagic fever (Sekamatte et al., 2018) by the Global Health Security Agenda (Kabami, 2023; Muturi et al., 2023). In some African countries like Sudan, Uganda, RVF has been reported as causing 10% mortality among adult livestock and abortions in ∼ 80-90% pregnant animals (Bird et al., 2009; Birungi et al., 2021; Hassan et al., 2020).

Uganda reported the isolation of the RVFV from mosquitoes in 1944 in Semliki Forest, Western Uganda (Dick, 1953). However, the first laboratory confirmed cases from animals and humans were not reported until 2016 from Kabale District (Shoemaker et al., 2019). Since then there have been sporadic RVF outbreaks in different parts of the country and these have been investigated by Ministry of Health (MoH) and Ministry of Agriculture, Animal Industries and Fisheries (MAAIF)’s surveillance departments (Aceng et al., 2023; Birungi et al., 2021; Kabami, 2023; Nyakarahuka et al., 2018a, 2023c, 2023a; Tumusiime et al., 2023a, 2023c). However, RVF surveillance is typically only carried out responsively after outbreaks that have already led to the loss of human lives or livestock. Proactive early warning surveillance systems based on known signs and symptoms, such as anomalous patterns of abortion in livestock, could help to minimize human RVF incidence and to promote livestock health (Gachohi et al., 2024).

The rate of abortion among pregnant ewes is reported to be almost 100% and over 90% of lambs infected with RVF die (WHO, 2022). Livestock abortions are a well-documented production and economic burden to famers, especially among livestock farming communities and countries that depend on agriculture (Gachohi et al., 2024; Kaur et al., 2023; Lokamar et al., 2020). In addition to the lost foetuses, abortion poses an additional excess mortality risk for female animals (Muma et al., 2006). Livestock owners lose the potential financial benefits that would be achieved through the sale of lost animals and animal products including beef, hides, skins and milk (Kabami, 2023; Muturi et al., 2023).

Budasha et al. conducted a random cross sectional study of livestock abortions in Kisoro district, Uganda (close to our study area in Isingiro) and reported an overall prevalence for livestock abortions of 17% with sheep having the highest prevalence of all livestock kept in the district (Budasha et al., 2018). Our study was motivated by Budasha et al. and de Glanville, W. A. et al.’s observation that RVF antibodies could be detected in abortions and their recommendation that these could be used as an early warning signal for RVF in endemic areas (de Glanville et al., 2022; Glanville et al., 2022). In Uganda and many African countries where animal health surveillance is not systematic (Hasahya et al., 2023; Qiu et al., 2023) livestock abortions are neither routinely documented nor reported for action, yet could be leveraged as cost-effective proxies for arboviruses surveillance in ruminants (Walt et al., 2023).

In this project we piloted a community led reporting system for abortions based on farmers self-reporting using phones. Reports were followed up by investigation and collection of samples to establish the potential exposure to RVF among recent cases of abortion (less than 14 days) using ELISA. ELISA kits are commonly used in low-resource settings (Domfe et al., 2022) and are recommended by the World Organization for Animal Health for the purpose of estimating the prevalence of infection of RVF (Petrova et al., 2020). A similar syndromic surveillance system piloted in rural settings of Kenya showed that animal health illness events were 15 times more likely to be reported by phone-based surveillance than home visits by veterinary health workers (Thumbi et al., 2019). Thumbi et al. recommended that a phone-based surveillance system could be implemented to enable the early detection of zoonotic diseases like RVF (Thumbi et al., 2019). In our project, we built on the methodology of Thumbi et al. adding publicity through social media channels, community sensitizations to increase awareness, and laboratory confirmation of reported and investigated abortions cases in livestock with a focus on RVF.

## Materials and Methods

### a) Study area

Isingiro district (0.84° S, 30.80° E) is located in southwestern Uganda, about 297 kilometres from the capital city, Kampala, and 47 km from Mbarara city. Isingiro shares a border with Tanzania in the south and three further districts of Uganda: Kiruhura, Rakai and Ntungamo (Fig 1). These districts are characterised by livestock rearing being the major industry on which residents depend for their livelihood (Adonia, 2013; Bwengye et al., 2023; Mubiru et al., 2023). The district also neighbors Lake Mburo National Game Park which harbours wildlife species susceptible to RVF. Both wildlife and livestock share common water sources, along the shores of Lake Mburo and the Kagera River which spans the district. Isingiro district has an estimated cattle, goats and sheep of population of 368,246, 422,108 and 88,621 respectively (UBOS, 2024) and human population of 486,360 based on projections from human 2014 census respectively (Tumusiime et al., 2023a;). Isingiro district has 35 lower administrative units (sub counties) with 18 where animal rearing is conducted. The district experiences a tropical savannah climate with an average annual rainfall of 1200mm, and a temperature range of 17 - 30°C (Kweyu et al., 2023; Nagasha et al., 2019). The district has two rainy seasons in each year: March to April, and September to November. The people in this area rear livestock and practice crop agriculture as their main economic activities (Taremwa et al., 2020).

**Figure 1:**
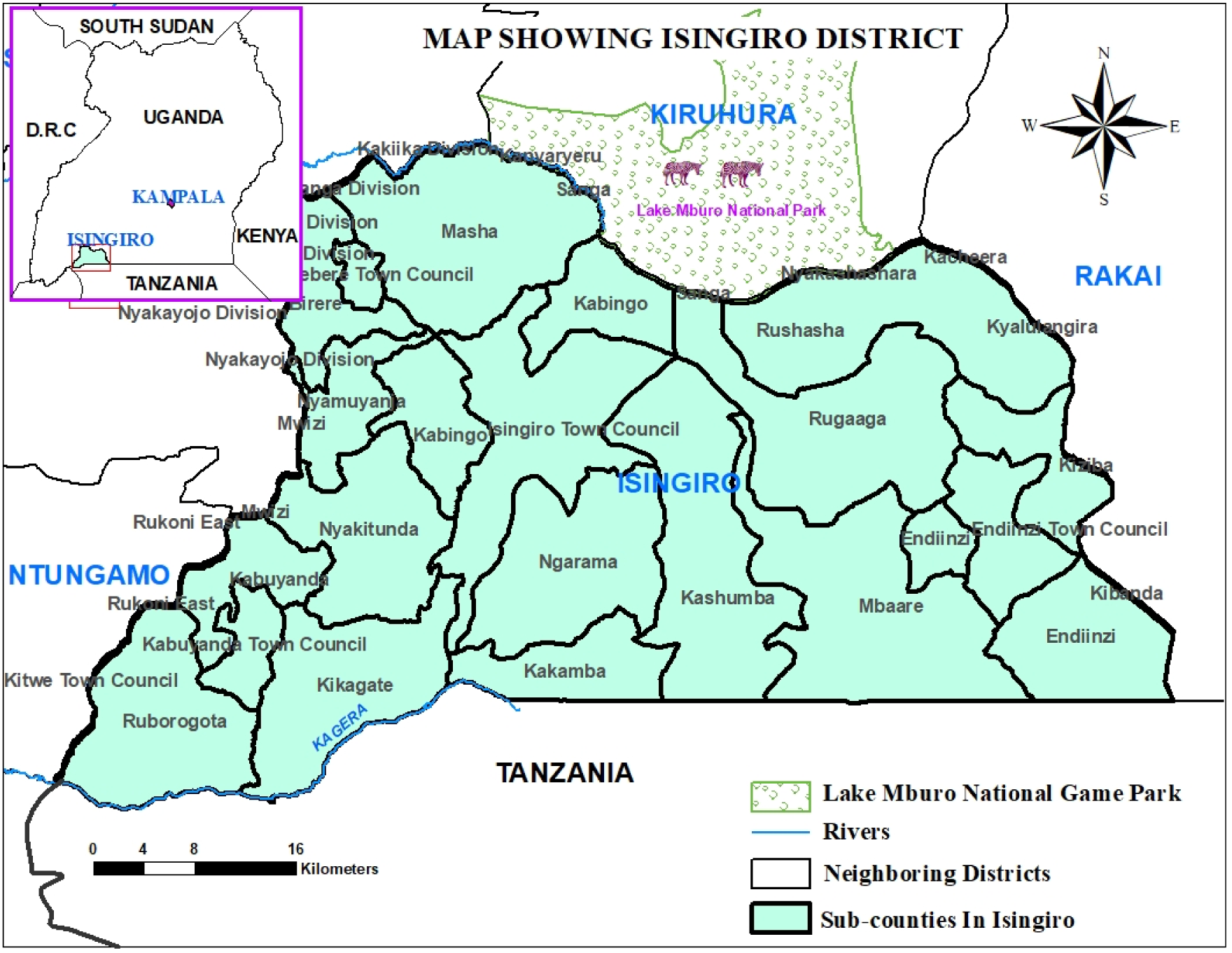
Map of Uganda showing Isingiro District and sites for sample collection.

### a) Study Design

We conducted a cross sectional study from March to June 2023. A call centre was established in consultation with livestock owners (farmers), local authorities and District Veterinary Officers (DVO), to receive and record reported livestock abortions. First, we carried out formal entry/information meetings to explain the purpose of the study, the information to be collected, how they were expected to participate and how to use the call centre. Further meetings were held with staff at the National Animal Diseases Diagnostics Epidemiological Centre (NADDEC), MAAIF and local veterinary caregivers briefing them on procedures for sample collection, packing and transportation to the laboratory in Entebbe for analysis.

The call centre was officially established in the office of the DVO at Isingiro district on 1^st^ March 2023 and co-managed by the corresponding author and a local contact officer who served as a liason person between this project and the DVO (Figure 2). Following the official set-up of the call centre, we were keen to follow-up abortion alerts that had occurred within the target 14 days period (14^th^ February 2023 onwards). However, we also considered and responded to abortion cases which had occurred before the set-up of the call centre. Establishment of the call centre was followed by different stakeholder engagements through meetings and mass sensitization of livestock owners and local leaders. Information Education and Communication (IEC) materials on RVF were created. These materials were then distributed to public gatherings, barazas and similar events to advocate for the reporting of abortions.

**Figure 2:**
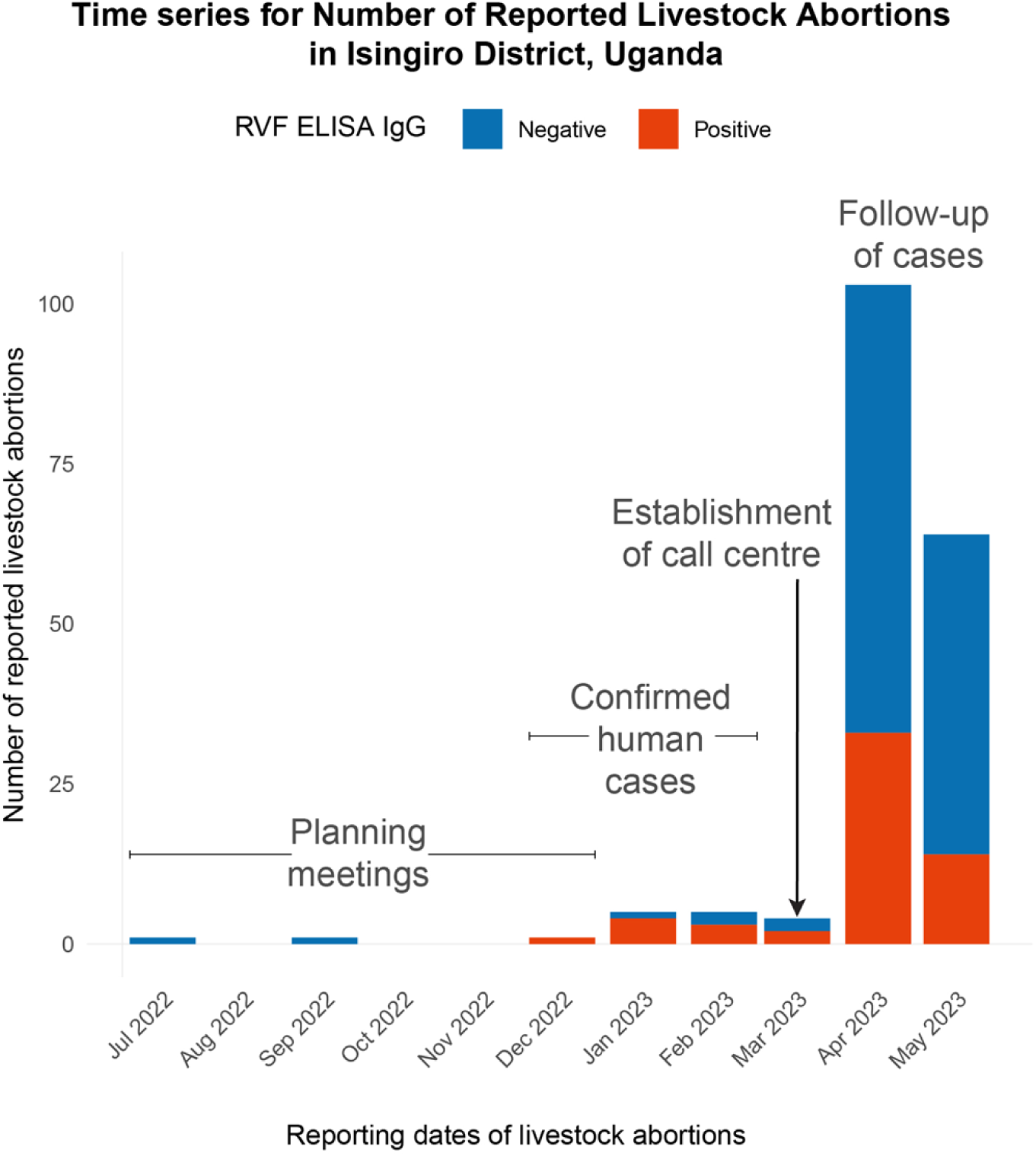
Time-series for livestock abortions reporting in Isingiro District, Uganda.

The contact officer was provided with a data collection tool, KoboCollect (Lakshminarasimhappa, 2022) installed on android phones which enabled tracking of all incoming calls regarding livestock abortions. At the design phase we expected that these phones could facilitate reporting through phone calls, SMS, and social media alerts (WhatsApp, Facebook) in addition to verbal reporting through walk-ins to the office. We set up two days’ of orientation meetings with different stakeholders including the research team that we had recruited. During these meetings, the participants were taken through the line-up of activities and their expected roles. This was necessary to ensure that stakeholders appreciated the scope of the project and would be more likely to support it. For the research team this was necessary as it helped them to understand the project protocol. On the last day of the orientation, consent was sought and a social media channel group [(WhatsApp)](https://chat.whatsapp.com/E08jaDD6u4MLTK9c356YY3) was formed. This was done to coordinate the reporting of livestock abortion reports across the district. High-level coordination of activities and events was carried out through a separate social media group (WhatsApp group) which included the corresponding author, District Health Officer (DHO), DVO, Assistant District Health Officer-Environmental Health (ADHO-EH), call centre staff and all research assistants. There were also weekly update meetings organised to review progress.

We provided two telephone numbers (the DVO and the contact officer) on materials to enable livestock abortions to continue to be reported after the end of the project. IEC materials in the form of flyers and brochures (in English, S2 Appendix and S3 Appendix) were adapted from RVF risk communication materials created by the Ugandan Ministry of Health. These materials were distributed and displayed in public places in the community and provided to local authority leaders for use in addressing the public during community/social events like burials, ceremonies, radio talk shows and community barazas. We also made announcements on local radio and carried out sensitizations with community health workers. To inform the necessary scale of the study we carried out a sample size calculation to ensure we could estimate the expected prevalence (𝑝) in the area to a reasonable precision (𝑑). We used the Kish-Leslie formula (𝑛 = 𝑧^2^𝑝𝑞𝑑^2^) where the desired confidence interval was taken to be 95% (5% significance level) and expected prevalence of RVF during abortions (effect size) was taken as 21% (De Glanville, Allan et al. 2022) which gave a target sample size of 325.

## Field data collection

To provide capacity building in the local area and in an attempt to build in sustainability for our project, we identified and recruited veterinary and laboratory staff employed in the Isingiro district into the project. Working with staff that were already employed in this study area was not only cost-effective but later proved beneficial as it enabled expedition of activities as the local staff understood the local geography which eased the logistics of data collection. Through this project, we brought on board both veterinary staff working in public and private practice. In preparation for fieldwork, these veterinary and laboratory staff were given safety training on laboratory Standard Operating Procedures (SOPs), use of Personal Protective Equipment (PPE), human handling and restraint of animals. These SOPs enabled staff to adhere to animal welfare principles, but also to ensure personnel safety and adherence to biosecurity measures to curtail spread of disease between farms. The team was also trained on the approved study protocol and data collection tools for two days.

The investigation team was provided with equipment and consumables for collecting sera and vaginal swabs from aborted livestock. We responded to 78% (329/423) of the reported alerts. This was intended to inspire farmers to report abortion alerts to the call centre. After every alert, there was an investigation by the corresponding author and the veterinary staff. During these investigations, a questionnaire was used to collect background information from the animal owners/farmers. The collected information included key demographic and epidemiological variables such as herd size, RVF vaccination history, alert channel used, environmental factors like stagnate water, presence of shrubs/forests, irrigation dams, vegetation cover, economic activities. The blood and vaginal swab samples collected were transported in ice-conditioned icepacks (approximately four hours after sample collection) to Rwenkubo Health Centre IV (central laboratory hub for Isingiro district) and were stored at -20°C. This laboratory received daily vaginal swab samples from the veterinary staff. We collected 200 sera samples and 269 vaginal swabs and transported to NADDEC for analysis, which were then tested for both RVF and Brucellosis. The samples were transported through the Ministry of Health accredited National Laboratory system (Hub) (Kiyaga et al., 2013) under cold chain conditions specified by the manufacturer to the central testing laboratory (NADDEC) (Monje et al., 2021). Due to budgetary constraints these have not yet been analysed.

### d) Laboratory analysis

Blood samples were tested by Enzyme-linked immunosorbent assay (ELISA) for anti-RVF and anti-Brucella antibodies using validated commercial kits. Brucella has a high prevalence in Uganda (Bugeza et al., 2023) compared to other infections associated with abortion storms (Akwongo and Kakooza, 2022; Aruho et al., 2021; Muma et al., 2006). For RVF we used the RVF competitive multi-species ELISA Kit (ID VET, Montpellier, France) (Petrova et al., 2020). Although vaccination against RVF could stimulate the production of specific antibodies producing a positive IgM test result (Matsiela et al., 2023) vaccine status of animals was verified with farmers with none reporting they had purchased or used vaccines for RVF. Brucella assays were done using IBL - America IgG-ELISA Kits (Minneapolis, MN) as described by Nyamota et al (Nyamota et al., 2023).

### e) Statistical Analysis

All data analyses were carried out using R statistical software version 4.3.1 (R Core Team (2023)., n.d.). Confidence intervals (CI) for the binomial proportion of apparent prevalence of exposure to RVF and Brucella were calculated using the Clopper-Pearson method as implemented in the “binomial” function of the “DescTools” package (Signorell A (2023), n.d.). We developed logistic regression models to identify factors associated with test-positivity for RVF. The small number (3/184) of IgM positive sera found precluded their use for this purpose, so we used IgG test status as the dependent variable. Following univariate screening for farm level risk factors (S1 Table) with a cutoff of p<0.2 for inclusion in a multivariable model, we developed a multivariable model. Known biological risk factors (stage of pregnancy, animal host, history of animal movements, environmental conditions) were also forced into the multivariable model regardless of the univariate significance (Nyakarahuka et al., 2023c). The associations were assessed by odds ratios, with a 5% level for statistical significance. Time series data were visualised using ggplot2 (Hadley Wickham, 2016) and the tidyverse packages (Hadley et al, 2019). We tested the logistic regression output for 174 samples collected during the target period (less than 14 days) and found minimal changes to the qualitative model results.

## Results

At the planning phase of the fieldwork (21^st^ January 2023), there were a small number (5) of reports of livestock abortions that had been recorded by the DVO’s office independent of our study (Fig 2, S2 Table). We received a total of 53 call alerts leading to 423 abortions reported during the pilot from 53 farms/herds owners and 184 of these alerts were investigated and for this paper we report 184 samples (S3 Table). The number of samples collected during the target period were 174 with 10 that occurred before the establishment of the call centre. The ten abortions that occurred before establishment of the call centre were all from cattle reported from Bugango Town Council (6/10) and Mbaare subcounty (4/10). The 4/10 and 6/10 were estimated to be at early stage and middle of pregnancy respectively (S2 Table). All of the reports were through telephone calls as opposed to social media platforms like facebook and WhatsApp. There was high interest in participating in this pilot and demand from farmers to understand the cause for the livestock abortions. This pilot had been planned to take place over six months but due to the high rate of reports, and delay in starting due to logistic factors, it was completed within three months (Fig 2). Regarding the delay between the date of reporting and investigation, the majority were in less than one day. However, the mean delay was skewed to the right due to few cases that were reported before setting up the call centre (S1 Appendix).

Of the collected samples from abortions, the greatest number 106/184 (58%) were from cattle, followed by goats 66/184 (36%). The majority of cattle were crossbred 25/52 (48%) and 22/52 (42%) were local breeds. There was no history of RVF vaccination reported by farmers and the majority (98%) did not report any history of cross-border or inter-district movement. The median number of livestock abortions reported per herd was (4.0; IQR 3.0). The majority of our reporters were male 158/184 (86%) and 159/184 (86%) described herdsman as their main occupation. The median household size was 10; IQR 4.25 with majority, 81/184 (44%) described with no formal education. The reported clinical presentation of animals at farm level included; sudden onset of abortion among pregnant animals, weakness, unsteady gait, mucopurulent nasal discharge, profuse fetid diarrhoea and high fever. The observed environmental and climatic factors near the livestock farms and homestead included; bushes, recent rainfall (less than 14 days), heavy forests and stagnant water. The reported abortions originated from 18/33 (61%) of the sub counties where livestock was reared (S1 Table).

The seroprevalence of RVF IgG in cattle was 38% [95%CI 29 – 47], 33% in sheep [95%CI 14 – 61] and 20% in goats [95%CI 12 – 31]. The IgM seroprevalence was lower, with 8% in sheep [95%CI 1 – 35] followed by cattle at 2% [95%CI 1 – 6]. With respect to brucellosis, goats had a seroprevalence of 36% [95%CI 25 – 49]; 16% of cattle were positive [95%CI 11 – 23] but no sheep tested positive (Table 1).

**Table 1:** Serological results for RVF and Brucellosis.

| **RVF IgG** | **Host** | **Apparent prevalence of exposure (%) (95% CI)** | **Negative** | **Positive** | **Total number of animals, n (%)** |
| --- | --- | --- | --- | --- | --- |
|  | Cattle | 38 (29 - 47) | 66 | 40 | 106 (58%) |
|  | Goats | 20 (12- 31) | 53 | 13 | 66 (36%) |
|  | Sheep | 33 (14 - 61) | 8 | 4 | 12 (6%) |
| **RVF IgM** | Cattle | 2 (1 - 6) | 104 | 2 | 106 (58%) |
|  | Goats | 0 | 66 | 0 | 66 (36%) |
|  | Sheep | 8 (1 - 35) | 11 | 1 | 12 (6%) |
| **IgG Brucellosis ELISA** | Cattle | 16 (11 - 23) | 106 | 20 | 125 (63%) |
|  | Goats | 36 (25 - 49) | 37 | 21 | 58 (29%) |
|  | Sheep | 0 | 16 | 0 | 16 (8%) |
| **Both IgG (RVF and Brucellosis)** | Cattle | 9 (5 - 15) | 54 | 9 | 106 (58%) |
|  | Goats | 2 (0 - 8) | 42 | 1 | 66 (36%) |
|  | Sheep | 25 (9 - 53) | 5 | 3 | 12 (6%) |

Sheep were the species most likely to be positive to both RVF and brucellosis tests with 25% [95% CI 9 – 53] positive to IgG and the Brucellosis IgG ELISA, followed by cattle at 9% [95% CI 5 – 15] and lastly goats at 2% (95%CI 0 - 8) (Table 1).

Only one risk factor (host species) was statistically significantly associated with RVF IgG status. Cattle had the highest risk of positivity than goats and sheep (OR = 2.9 [95%CI: 1.27 - 7.07], p = 0.014). The presence of any favourable conditions (*Bushes, Recent rainfall (less than 14 days), Heavy forests, Stagnant water)* near the farm/herd (OR = 1.5 [95%CI: 0.6 - 4.0], p = 0.4) and history of movement (OR = 3.0 [95%CI: 0.1 - 71.0], p = 0.5) were not statistically significant in our sample despite their previously reported associations with risk of exposure to RVF (Table 2). As a sensitivity analysis, we refitted the multivariate model excluding the small number (10) of reports where the delay between alert and investigation was greater than 14 days. The qualitative results in terms of identified risk factors and magnitude of effects were unchanged for the smaller data set.

**Table 2:** Multivariate regression mode output for IgG RVF occurrence and associated risk factors.

| Variable | Number<br>n | Estimate | Std Error | Z value | OR<sup>1</sup> | 95% CI<sup>1</sup> | p-value |
| --- | --- | --- | --- | --- | --- | --- | --- |
| **Animal Hosts** |  |  |  |  |  |  |  |
| Goats | 66 | REF | - | - | - | - | - |
| Cattle | 106 | -1.07 | 0.44 | 2.45 | 2.9 | 1.3, 7.1 | **0.014\*** |
| Sheep | 12 | 1.012 | 0.73 | 1.39 | 2.8 | 0.6, 11.3 | 0.2 |
| **Stage of animal pregnancy** |  |  |  |  |  |  |  |
| Early stage<br>(1-3 months) | 73 | REF | - | - | - | - | - |
| Middle stage (4-6 months) | 62 | -0.74 | 0.48 | -1.51 | 0.5 | 0.18, 1.2 | 0.13 |
| Late stage (7-9 months) | 45 | -0.56 | 0.41 | - 1.37 | 0.6 | 0.3, 1.3 | 0.20 |
| Not sure | 4 | -16.77 | 1181.54 | - 0.01 | 0.0 | - | 1.0 |
| **Environmental features at the farm/herd level** |  |  |  |  |  |  |  |
| A** | 81 | REF | - | - | - | - | - |
| B** | 52 | 0.34 | 0.41 | 0.83 | 1.4 | 0.6, 3.2 | 0.41 |
| C** | 15 | 0.19 | 0.74 | 0.25 | 1.2 | 0.2, 4.8 | 0.80 |
| D** | 36 | 0.43 | 0.48 | 0.90 | 1.5 | 0.6, 4.0 | 0.37 |
| **History of Animal movement** |  |  |  |  |  |  |  |
| No | 181 | REF | - | - | - | - | - |
| Yes | 3 | 0.96 | 1.47 | 0.65 | 2.6 | 0.1, 71.0 | 0.52 |
| <sup>1</sup> OR = Odds Ratio, CI = Confidence Interval |  |  |  |  |  |  |  |

*\*Statistically significant variable*
**\*\*A:** Bushes, Recent rainfall (less than 14 days), Heavy forests, Stagnant water. **B:** Only 03 present; Bushes, Stagnant water, Heavy forests. **C:** Only two present; Bushes, Recent rainfall (less than 14 days). **D:** Only one present.

## Discussion

This study is the first of its kind in Uganda, testing a community-led system for reporting livestock abortions to detect Rift Valley Fever (RVF). After setting up a call center in Isingiro district, most incidences of abortion were reported, recorded in the log book and cattle showed the highest RVF prevalence, followed by sheep and then goats. Regarding brucellosis, goats had the highest prevalence than cattle. These findings align with previous research conducted in Uganda and Keny (Birungi et al., 2021; Kabami, 2023; Ndumu et al., 2021; Nyakarahuka et al., 2018a, 2023a, 2023b, 2023c; Nyamota et al., 2023; Tumusiime et al., 2023b).

Through this pilot we have demonstrated the feasibility of setting up a call centre for abortions. Such call centres are particularly valuable for animal disease surveillance in settings like Uganda where the majority of livestock farmers have phones but do not necessarily have internet access. When a similar pilot study was carried out in Kenya the authors recommended pathogen specific abortions reporting and investigation which our work has contributed to (Gachohi et al., 2024). Such call centres can provide added value to both farmers and policy makers. For example, farmers can seek technical guidance on farm management and have the opportunity to report any unusual or suspected conditions at the farm. At a policy level, they can complement existing surveillance infrastructure at Uganda’s Ministry of health and office of the Prime Minister such as the national level emergence operations centre (Ario et al., 2022) and the National One Health Platform (Buregyeya et al., 2020) respectively. These organisations have technical staff who can analyse and interpret phone reports. Furthermore, from the animal health sector perspective, the set up of a national level abortions surveillance system could build on existing systems like the electronic infectious diseases surveillance (eIDS) system under MAAIF. Looking beyond Uganda, neighbouring Kenya has already established such an integrated animal health system (Njenga et al., 2021). Setting up call centres has been previously championed under the Ministry of Health Uganda during both peak moments of different epidemics such as the SARS-CoV2 pandemic (Katana et al., 2024), for Ebola Virus disease (Kiggundu, 2022) and for longer-term endemic infections such as HIV/AIDS and Tuberculosis. Establishing district-level call centres in DVO offices to faclilitate livestock abortions reporting could proivide a cost-effective and sustainable path to improve animal health surveillance in Uganda.

Syndromic surveillance is cheap as most people/farmers have phones, although the follow-up/investigation of such reported cases may be expensive and require government investment. Investment in development of pen-side diagnostics for RVF or by expanding access to existing diagnostics through commercial or social enterprises could play an important role in addressing this limitation. However, the high costs of investigating reported cases could also be minimised by prioritising investigation of anomalous reports of abortions (either seasonally or geographically). Establishing such criteria will require a longer, and more geographically representative study than was possible for this pilot. Syndromic surveillance based on livestock abortions may be imprecise in estimating the risk of RVF, due to the other circulating diseases like brucellosis and Q-fever which will contribute to abortions, but is likely to be cheaper than entomological surveillance. We found a high demand and interest from farmers to participate in an abortion surveillance programme as they were eager to know the causes. This interest could be leveraged to promote community level reporting through call centres. The access and utilization of such point-of-care diagnostics would be built on grounds that farmers are willing to invest in diagnosing and treating their sick animals.

Our results suggest that RVF was circulating among livestock within these communities as evidenced by high IgG positivity across all livestock species. The relatively lower IgM positivity is suggestive of less current circulation despite the confirmed presence of RVF in four districts of Isingiro, Mbarara, Rubanda and Kazo in the 16^th^ – 20^th^ epidemiological weeks (Kabami, 2023). The presence of RVF antibodies in Isingiro district could also be explained by previous outbreaks in 2018, 2020 (Nyakarahuka et al., 2023c). The nationwide seroprevalence study by Nyakaruhuka et al showed that RVF was circulating in over 37/135 districts across the five regions in Uganda (Nyakarahuka et al., 2023c). An ecological niche model for Uganda also suggested that Isingiro district was a high risk area and the authors recommended surveillance activities to be targeted in such areas (Tumusiime et al., 2023a). In comparison with Kenya’s IgG RVF cattle seroprevalence picture (an indication to previous exposure to RVF), Owange et al reported 13.1% (183/1396) in 2014 (Owange et al., 2014a), Nanyingi et al reported 33.3% (95% CI [6.7–60]) in 2017, Bett et al reported 37.10% (30.72 – 43.84) in 2018. Our IgG seroprevalence of 38% is not dissimilar to that seen in endemically infected populations, however our study population of animals with recent abortion might be expected to be at a higher risk of exposure.

In our study, abortions were reported across different stages of pregnancy, ranging from early to late stages, which would be highly suggestive of endemic RVF (Oymans et al., 2020). According to the World Organization of Animal Health differential diagnosis of livestock diseases, RVF is expected to cause abortion storms (for epidemic) conditions but under endemic conditions it repeatedly causes abortions at different stages of pregnancy (Gerdes, 2004), as reported here.

We found animals with both RVF and brucellosis antibodies. Given that our entry point was livestock abortions to confirm potential causes, our screening for RVF and brucellosis was important as both are likely to contribute to the occurrence of abortions. This finding underscores the need to set up integrated livestock diseases surveillance which could be supported by development and deployment of point-of-care diagnostics that have multiple assays. Early warning systems could be designed with different signals to distinguish the contribution of RVF to observed abortions in comparison to other abortion causing pathogens. Cattle had a higher risk to be positive for IgG antibodies for RVF. This observed higher risk of exposure in cattle may simply be done to the longer expected lifespan of this species compared to sheep and goats. This longer duration could be linked to thus the IgG antibodies. Several studies have reported similar findings, linking higher prevalence in cattle to the greater opportunity for exposure to RVF virus (Jeanmaire et al., 2011; Nyakarahuka et al., 2018b; Owange et al., 2014b). However, this observation is quite different from the historically known RVF epidemiology which implicates sheep and goats more than cattle (Anywaine et al., 2022; Birungi et al., 2021). This discrepancy could potentially be explained by the far higher number of cattle in our study and the distinction between risk of clinical illness and the presence of IgG antibodies which we measure.

Cattle contributed the highest number of samples collected followed by goats and sheep. Multiple reports/samples taken from the same farm could have skewed distribution towards cattle, or it may be that farmers were more likely to report cattle due to their value. Previous studies in Uganda (Nyakarahuka et al., 2018, 2023a, 2023c) and neighbouring countries like Kenya (Hassan et al., 2020; Muturi et al., 2023) have also found surprisingly low rates of reports of abortions in sheep. However, the small number of sheep included in the study, means we could not measure the (relative) risk of infection infection in this species. The small number of sheep in the study is a representative of national-wide livestock statistics picture, for example the 2021 livestock census showed that Uganda had 17.4 million goats, 14.5 Million cattle and 4.4 million sheep (UBOS, 2024).

The risk of introduction of RVF through livestock movements has also been highlighted by previous studies (Kim et al., 2021; Tigoi et al., 2020). Despite hearing anecodotal reports of cross-border movements between Uganda and Tanzania throughout Isingiro District through the porous borders and nomadic grazing grounds, livestock movement was not a statistically significant factor in our study. This could be a consequence of our sample size but most likely the fewer number of farms reporting livestock movements. For example only 3/184 farms reported history of animal movements. The tracking of animal movements in Uganda is not well established and synchronized (González-Gordon et al., 2023; Hasahya et al., 2023; Payne et al., 2021). For example, the available animal movement tracking efforts are for those to be transported on major/highway roads and the motivation is trade and revenue generation. The movements at sub-national/districts level like our study area is hardly tracked yet such data would be very important. Even with the minimal animal movements reported could not be verified at farm level by the research team. In addition, there is poor book keeping at farm level in our study area. Therefore, there should be caution in interpreting our model output on this particular variable. We recommend an intentional animal movement registry at farm level that could sychnronize with a national-level database for animal movements. This is specifically important at farm level because the few national-level tracked animal movements 34% are for breeding purposes yet this movement poses a risk for disease transmission between farms.

We had expected that social media platforms would have enhanced the ease of reporting but found that telephone was overwhelmingly the preferred mode of reporting livestock abortions. We attribute this to the rural nature of the study area that is faced with challenges for electricity and internet connectivity which jeopadrises the use of internet-based/online based reporting systems (J et al., 2023; Lo and Kibalya, 2023). Our results therefore suggest that mobile phones are still the most effective platform for the establishment of syndromic surveillance system in low-income countries where internet connectivity is not well developed.

Our study area was characterized by environmental features such as shrubs and grass, areas of stagnant water and the slow flow of River Kagera all of which are likely to contribute to survival and breeding of the Culex and Aedes mosquito species which are critical for RVF spread (Muturi et al., 2023; Nanyingi et al., 2015). Given the ubiquity of these features in the study area it is perhaps not surprising that environmental factors were not identified as risk factors.

Our study had a number of strengths; we have piloted the establishment of livestock abortions reporting/surveillance and showed that this is feasible. We have also demonstrated that investigation of livestock abortions can aid diagnosis of brucellosis and RVF. We have added a further justification on the prioritisation of phone-based reporting for community level disease surveillance in low-income countries where internet/online platforms are not well developed. We innovatively recruited private veterinary officers/practitioners who sell veterinary services like drugs to the livestock farmers. The integration of private practitioners in surveillance could be adopted by MAAIF to help meet the most pressing animal health needs of the community. We developed RVF IEC materials and built capacity for public health and veterinary staff working with Isingiro district which is very important for sustainability of livestock abortions surveillance.

There were some limitations in our study; we conducted this study over a relatively short period (March – June 2023) and could therefore not measure any seasonal patterns in livestock abortions. The high demand from farmers mean that we exhausted our resources for testing in only three months (rather than the six months originally planned). Better understanding of the seasonal pattern of abortions would help the development of early warning systems for RVF given the strong seasonal risk of exposure driven by mosquito population dynamics. The demand and willingness for farmers to participate in this pilot study demonstrates the feasibility of carrying out a longer study in these communities. Extending our study over (at least) a year and a wider geographic range could allow the association between abortion rates and RVF transmission to be modelled and potentially to identify early warning signals that do not depend on expensive diagnostics.

We only sampled animals with evidence of recent abortions denying us the opportunity to assess the risk of RVF in the general livestock population. Being a cross-sectional study, we cannot test for a causal relationship between livestock abortions and RVF infection. Due to budget constraints, we only screened for RVF and Brucellosis, yet there are other pathogens (parasitic, fungal, viral and bacterial) which are also known to cause abortions in livestock (in particular bluetongue virus, Q fever and *Campylobacter* spp.). Ticks are common in the study area which could contribute to abortions from other causes.

## Conclusion

Through this study, we established the feasibility of setting up call centres for surveillance of livestock abortions in Uganda and found a high prevelance of exposure to RVF in animals that have recently experienced abortions. The demand and interest of livestock farmers to participate in such a surveillance programme was high as they are eager to know the cause(s) of abortions. Despite this, reporting may only be sustained if reported calls result in follow-up investigations and results disseminated to the farmers. This being a cross sectional study, it is not possible to infer a causal association between RVF and abortions but our estimates of seroprevalence are similar to those in neighbouring countries like Kenya where RVF is endemic.

## Data Availability

The data that support the findings of this study are publicly available from (https://github.com/abelwalekhwa/Livestock_Abortions) with the identifier(s) as datasets, R code.

## Acknowledgments

First, we acknowledge technical guidance and permission to conduct this study from Dr. Anna Rose Ademun, the commissioner Animal Health – MAAIF. We are grateful for funding for this fieldwork from Biotechnology and Biological Sciences Research Council (BBSRC) Impact Acceleration Account award, Public Engagement Starter Grant, Nigeria Travel Grant from University of Cambridge Centre from African studies. AJKC and JLNW were supported by The Alborada Trust. AWW was supported by Cambridge Trust for his doctoral studies at University of Cambridge, United Kingdom. Furthermore, special appreciation to CEHA for administrative support during this fieldwork in Uganda.

We acknowledge Dr Robert Ofwete and Micheal Wambi for their support in spatial visualization of Fig 1 and 2 respectively. We acknowledge the farmers whom we worked closely during the establishment of this pilot project in Isingiro District, Uganda. The technical guidance during initial stages of conceptualization of this idea from Drs Jennifer Lord and Joshua Longbottom are highly appreciated. The incredible grateful support from Ministry of Agriculture, Animal Industries and Fisheries and Isingiro District Local Government for my fieldwork in Uganda. Specifically, Isingiro staff including Dr Bruhan Kasozi (District Veterinary Officer), Dr Edson Tumusherure (District Health Officer), Marion Alowo, Pius Manigaruhanga, entire district health team and veterinary staff. MAAIF staff; Dr Dan Tumusiime (Senior Veterinary Officer), Dr Ben Ssenkera (Senior Veterinary Officer), Dr Robert Mwebe (Principal Veterinary Officer), Mr Olympia Mugarura and Mr Milton Bahati, both laboratory technologist. Lastly, we appreciate the contribution of different research assistants led by Mellon Ainembabazi and Noel Emma Esutu.

## Ethical Considerations

This study is part of the corresponding authors PhD project. We sought and received ethical approval to conduct the research from Makerere University School of Public Health (Reference No: *SPH-2022-364)*, Uganda National Council of Science and Technology (UNCST) (Reference No: *A264ES),* and Human Biology Research Ethics Committee, University of Cambridge, United Kingdom (Reference No: *HBREC.2023.02*). Lastly, we sought administrative clearance from Isingiro district local governments’ administration specifically from the Chief Administrative Officer’s office (dated *20^th^ March 2023*) for this study to be conducted.

Written informed consent from the research participants (Farmers/livestock owners) was obtained for free and voluntary participation in the study. This was after understanding the purpose and costs of the study. During the blood and vaginal swab collection, we also sought written consent for photographing of the animals for potential detection of RVF and follow-up for actions in future. These photos were only for identification of the animals should the vaginal swabs turn positive for RVF for control strategies by Isingiro district Veterinary department or MAAIF. To enhance understanding, we translated all the data collection tools and informed consent into the local language (Runyankole) which was the most spoken language in the Isingiro district. This will ease the understanding of the entire project for the participants to consent. We further kept the information confidential by keeping identifying information (telephone numbers) under key and lock but also coded. The data was analysed and published in aggregate form to avoid identification of individual participants. The data is currently stored under key and lock for the five years after publication as stipulated by UNCST.

## Author Contributions

AWW, AJKC and JLNW conceptualized the study, developed protocols and analysed the data. AJKC mobilized funds to implement this pilot study, LM supervised AWW during his fieldwork in Uganda. EH, ARA and SAA gave technical guidance to AWW during fieldwork in Isingiro district and laboratory analysis at NADDEC. AWW and AJKC drafted the first version of the manuscript. EH, LM, SAA, ARA and JLNW reviewed and gave technical input to the manuscript. All authors approved the final version of the manuscript.

## Supporting information

**S1 Table:** Socio-demographic characteristics of respondents.

| **Variable** | **Frequency (n)** | **Percentages (%)** |
| --- | --- | --- |
| **Animal species tested for RVF** |  |  |
| Cattle | 106 | 58 |
| Goats | 66 | 36 |
| Sheep | 12 | 7 |
| **Animal breeds in the study area (n=52)** |  |  |
| Exotic | 5 | 10 |
| Crossbreed/hybrid | 25 | 48 |
| Locals (Ankole (*Bos taurus indicus*), East African shorthorn Zebu (*Bos indicus*)) | 22 | 42 |
| **Gender of respondent** |  |  |
| Females | 26 | 14 |
| Males | 158 | 86 |
| **Means of reporting to the call centre (n=52)** |  |  |
| Phone Calls | 52 | 100 |
| **Number of livestock abortions reported (n=52)** |  |  |
| Median (4.0 IQR 3.0) |  |  |
| **Occupation of the respondent** |  |  |
| Transport cyclist | 2 | 1 |
| Teacher | 6 | 3 |
| Small scale farmers | 5 | 3 |
| Herdsmen | 159 | 86 |
| Political leaders | 5 | 3 |
| Others (religious leader, soldier, business) | 4 | 2 |
| **Education level of the respondent** |  |  |
| Advanced level | 14 | 8 |
| Bachelors | 5 | 3 |
| Certificate | 13 | 7 |
| Diploma | 12 | 7 |
| No education | 81 | 44 |
| Ordinary level | 58 | 32 |
| Masters degree | 1 | 0 |
| **Household size** |  |  |
| Median; 10 IQR 4.25 |  |  |
| **Animal Clinical presentations at farm level\*** |  |  |
| W | 95 | 52 |
| X | 29 | 16 |
| Y | 18 | 10 |
| Z | 24 | 13 |
| I don't know | 18 | 10 |
| **Stages of the pregnancy** |  |  |
| Early stage ( 1-3 months) | 73 | 40 |
| Middle stage (4-6 months) | 62 | 34 |
| Late stage (7-9 months) | 45 | 24 |
| Not sure | 4 | 2 |
| **Environmental Conditions around the farm\*\*** |  |  |
| A | 81 | 44 |
| B | 52 | 28 |
| C | 15 | 8 |
| D | 36 | 20 |
| **History of animal movement** |  |  |
| No | 181 | 98 |
| Yes | 3 | 2 |
| **Reported Animal vaccination status** |  |  |
| No | 184 | 100 |
| **Total** | **184** | **100** |

*\*W: Sudden onset of abortion among pregnant animals, Weakness /Unsteady gait Mucopurulent nasal discharge, Profuse fetid diarrhoea, High fever. X: Any three of the above symptoms, Y: Any two of the above symptoms, Z: Any one of the above symptoms*
*\*\* A: Bushes, Recent rainfall (less than 14 days), Heavy forests, Stagnant water. B: Only 3 present; Bushes, Stagnant water, Heavy forests. C: Only two present; Bushes, Recent rainfall (less than 14 days). D: Only one present.*

**S2 Table:**
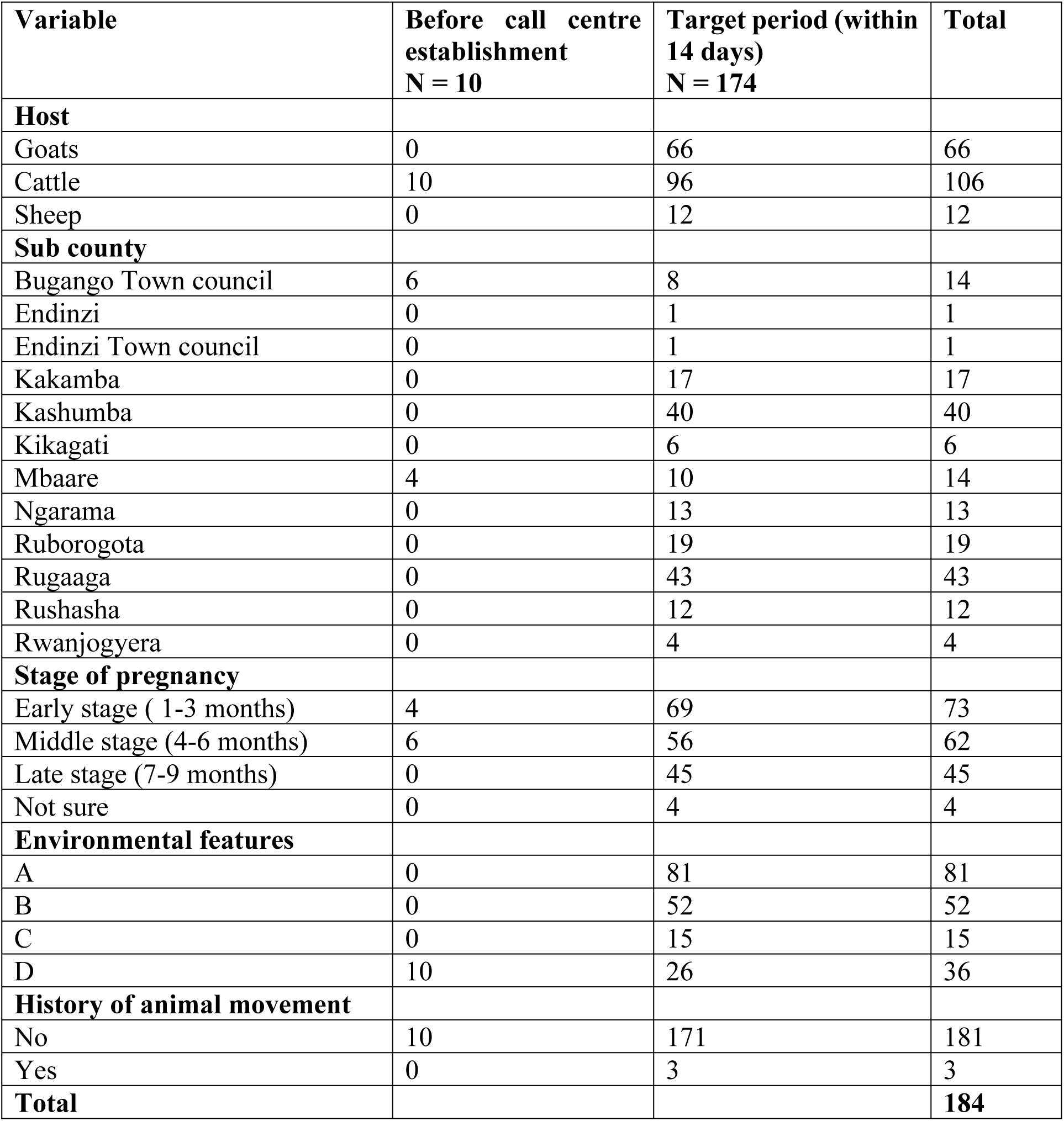
Period for reporting of abortion cases.

| **Variable** | **Before call centre establishment**<br>**N = 10** | **Target period (within 14 days)**<br>**N = 174** | **Total** |
| --- | --- | --- | --- |
| **Host** |  |  |  |
| Goats | 0 | 66 | 66 |
| Cattle | 10 | 96 | 106 |
| Sheep | 0 | 12 | 12 |
| **Sub county** |  |  |  |
| Bugango Town council | 6 | 8 | 14 |
| Endinzi | 0 | 1 | 1 |
| Endinzi Town council | 0 | 1 | 1 |
| Kakamba | 0 | 17 | 17 |
| Kashumba | 0 | 40 | 40 |
| Kikagati | 0 | 6 | 6 |
| Mbaare | 4 | 10 | 14 |
| Ngarama | 0 | 13 | 13 |
| Ruborogota | 0 | 19 | 19 |
| Rugaaga | 0 | 43 | 43 |
| Rushasha | 0 | 12 | 12 |
| Rwanjogyera | 0 | 4 | 4 |
| **Stage of pregnancy** |  |  |  |
| Early stage ( 1-3 months) | 4 | 69 | 73 |
| Middle stage (4-6 months) | 6 | 56 | 62 |
| Late stage (7-9 months) | 0 | 45 | 45 |
| Not sure | 0 | 4 | 4 |
| **Environmental features** |  |  |  |
| A | 0 | 81 | 81 |
| B | 0 | 52 | 52 |
| C | 0 | 15 | 15 |
| D | 10 | 26 | 36 |
| **History of animal movement** |  |  |  |
| No | 10 | 171 | 181 |
| Yes | 0 | 3 | 3 |
| **Total** |  |  | **184** |

**S3 Table:** Total number of abortion cases reported at the call centre.

| Call centre Alert # | Geographical locations (subcounties) | Number of reported abortions (N) | Abortion cases investigated/ Sera samples collected (n) | Proportion sampled (%) |
| --- | --- | --- | --- | --- |
| 1 | Masha | 3 | * |  |
| 2 | Masha | 1 | * |  |
| 3 | Rwentango | 10 | * |  |
| 4 | Ngarama | 21 | 13 | 57 |
| 5 | Ngarama | 2 |  |  |
| 6 | Kashumba | 4 | 40 | 66 |
| 7 | Kashumba | 3 |  |  |
| 8 | Kashumba | 1 |  |  |
| 9 | Kashumba | 30 |  |  |
| 10 | Kashumba | 3 |  |  |
| 11 | Kashumba | 5 |  |  |
| 12 | Kashumba | 4 |  |  |
| 13 | Kashumba | 8 |  |  |
| 14 | Kashumba | 3 |  |  |
| 15 | Bugango | 2 | * |  |
| 16 | Bugango T/C | 6 | 14 | 100 |
| 17 | Bugango T/C | 3 |  |  |
| 50 | Bugango T/C | 5 |  |  |
| 18 | Mbaare | 10 | 14 | 78 |
| 19 | Mbaare | 8 |  |  |
| 20 | Nakivale | 1 | * |  |
| 21 | Nakivale | 2 | * |  |
| 22 | Nakivale | 1 | * |  |
| 23 | Nakivale | 2 | * |  |
| 24 | Kakamba | 24 | 17 | 71 |
| 25 | Rushasha | 28 | 12 | 43 |
| 26 | Endizi T/C | 10 | 1 | 2 |
| 27 | Endizi T/C | 8 |  |  |
| 28 | Endizi T/C | 6 |  |  |
| 29 | Endizi T/C | 5 |  |  |
| 30 | Endizi T/C | 6 |  |  |
| 31 | Endizi T/C | 5 |  |  |
| 32 | Endizi T/C | 3 |  |  |
| 35 | Endizi T/C | 5 |  |  |
| 36 | Endizi T/C | 14 |  |  |
| 37 | Endizi T/C | 5 |  |  |
| 38 | Endizi T/C | 4 |  |  |
| 39 | Endizi T/C | 3 |  |  |
| 40 | Endizi Subcounty | 16 | 1 | 3 |
| 41 | Endizi Subcounty | 3 |  |  |
| 42 | Endizi Subcounty | 8 |  |  |
| 43 | Endizi Subcounty | 4 |  |  |
| 44 | Endizi Subcounty | 3 |  |  |
| 45 | Endizi Subcounty | 6 |  |  |
| 46 | Rugaga | 9 | 43 | 72 |
| 47 | Rugaga | 25 |  |  |
| 48 | Rugaga | 23 |  |  |
| 49 | Rugaga | 3 |  |  |
| 51 | Ruborogota | 22 | 19 | 86 |
| 52 | Kikagati | 30 | 6 | 19 |
| 53 | Kikagati | 2 |  |  |
| 33 | Rwanjogyera | 4 | 4 | 80 |
| 34 | Rwanjogyera | 1 |  |  |
|  | **Total** | **423** | **184** |  |

*\*Lost samples due to labelling loss during transportation to the national laboratory*

**S1 Appendix:**
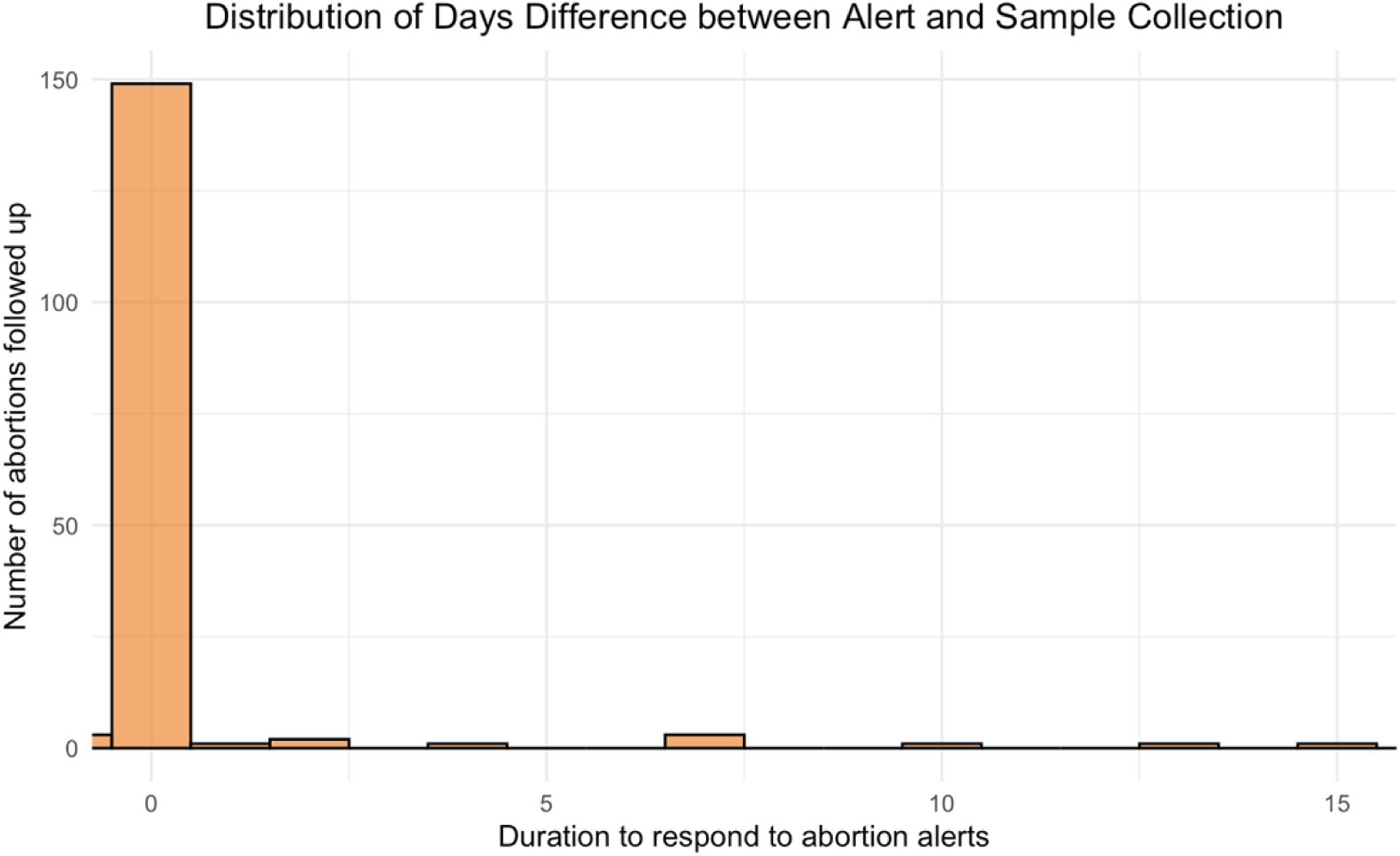
Delay between alert and investigation (days)

**S2 Appendix:**
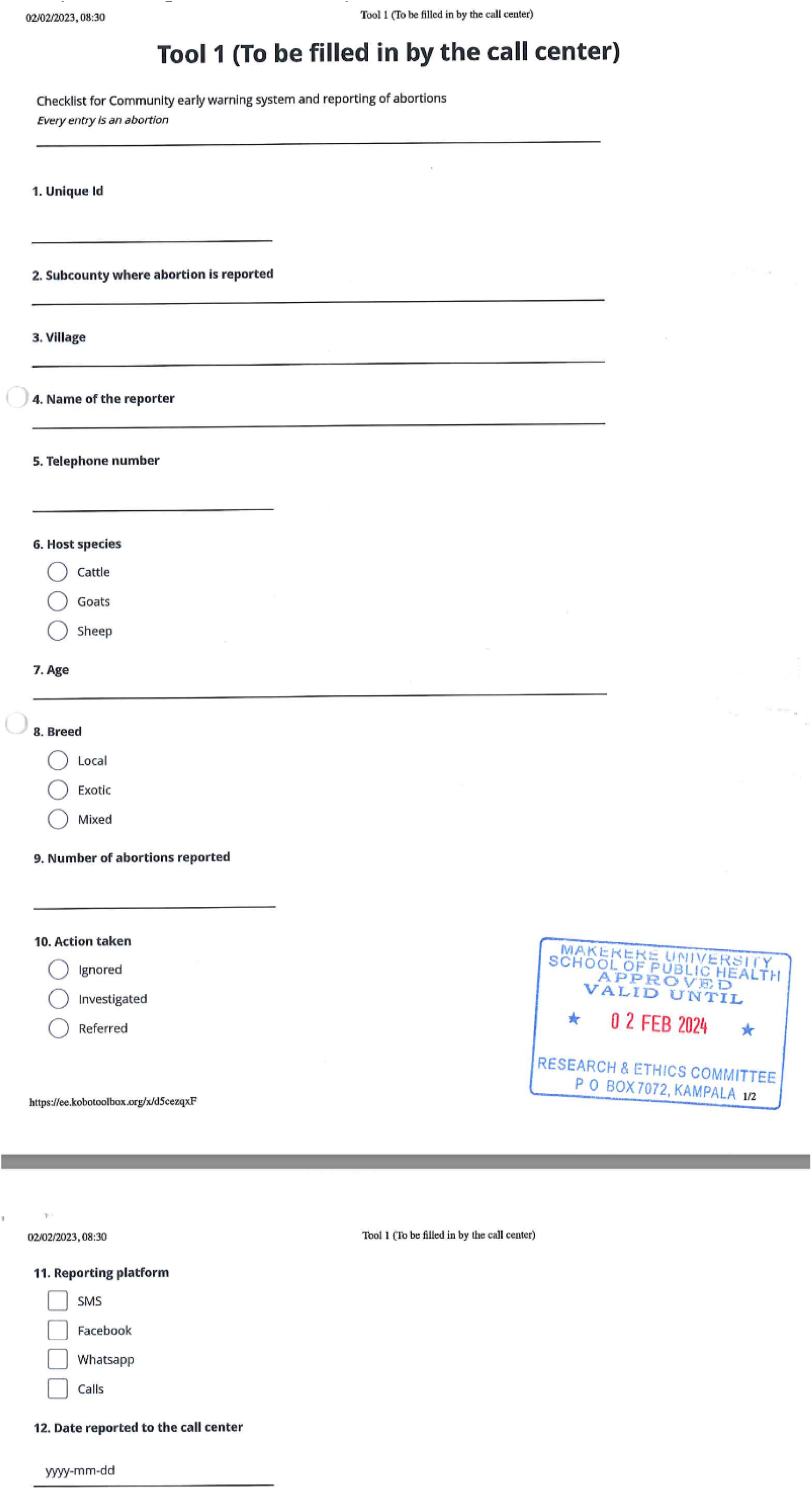
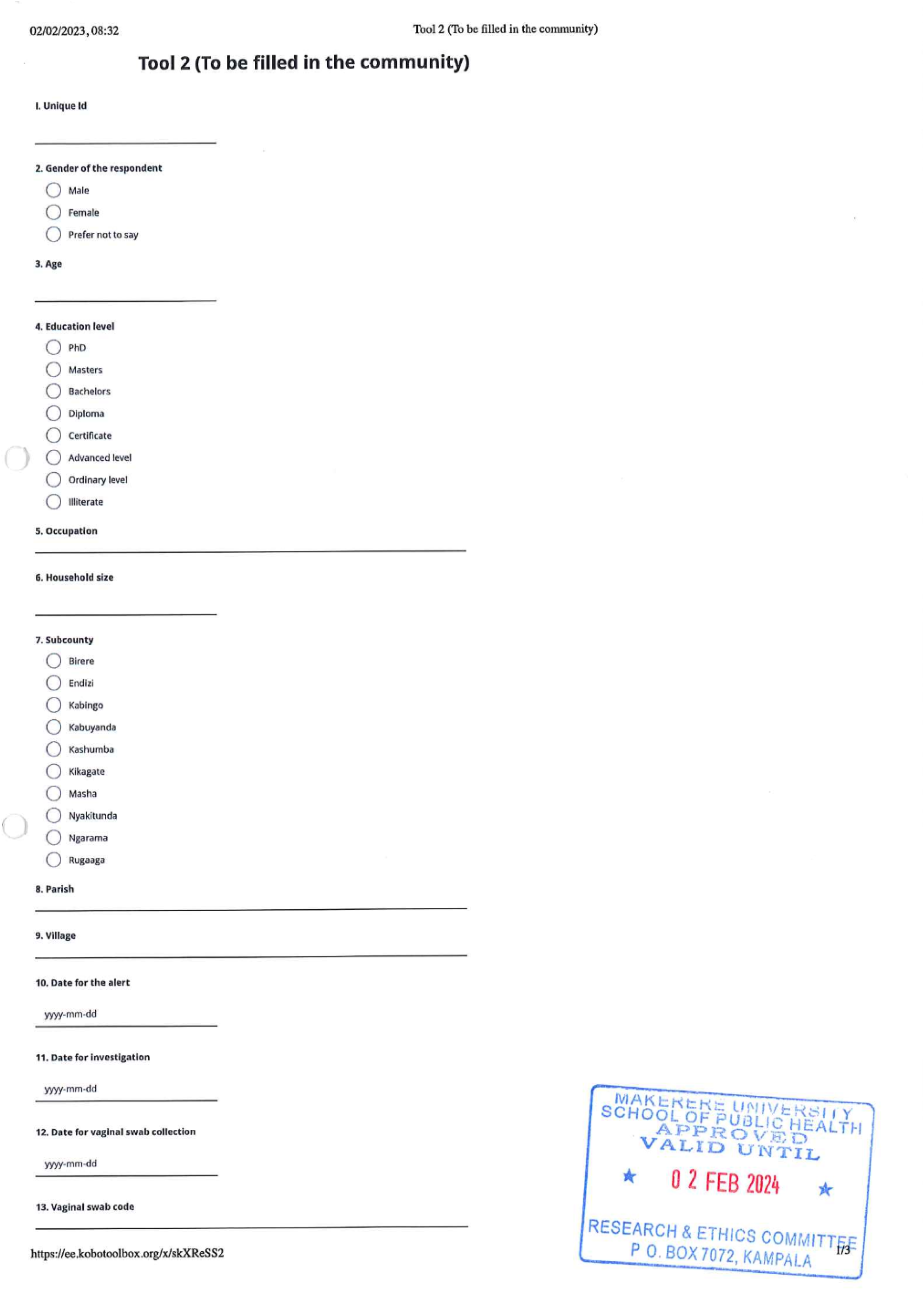
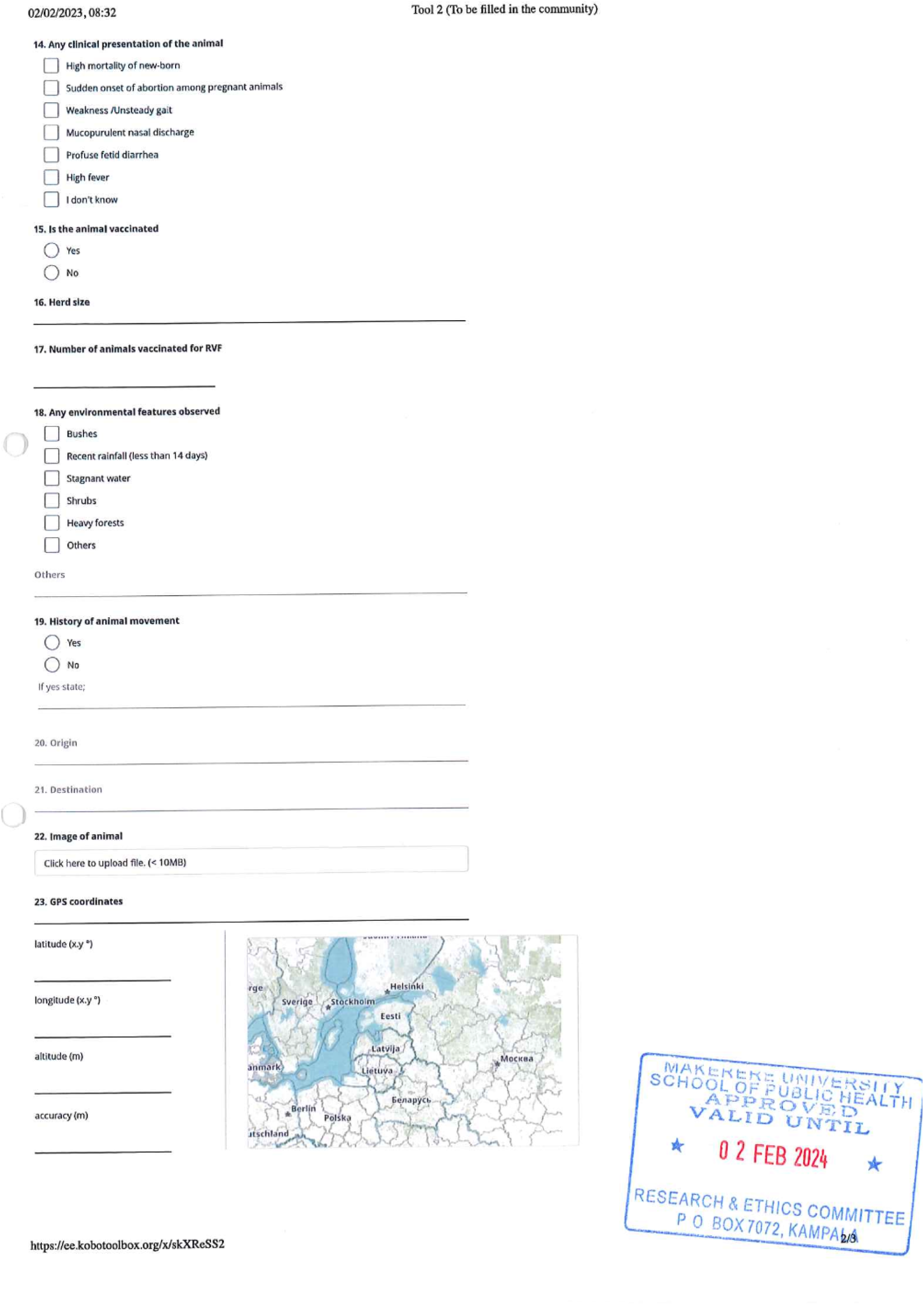
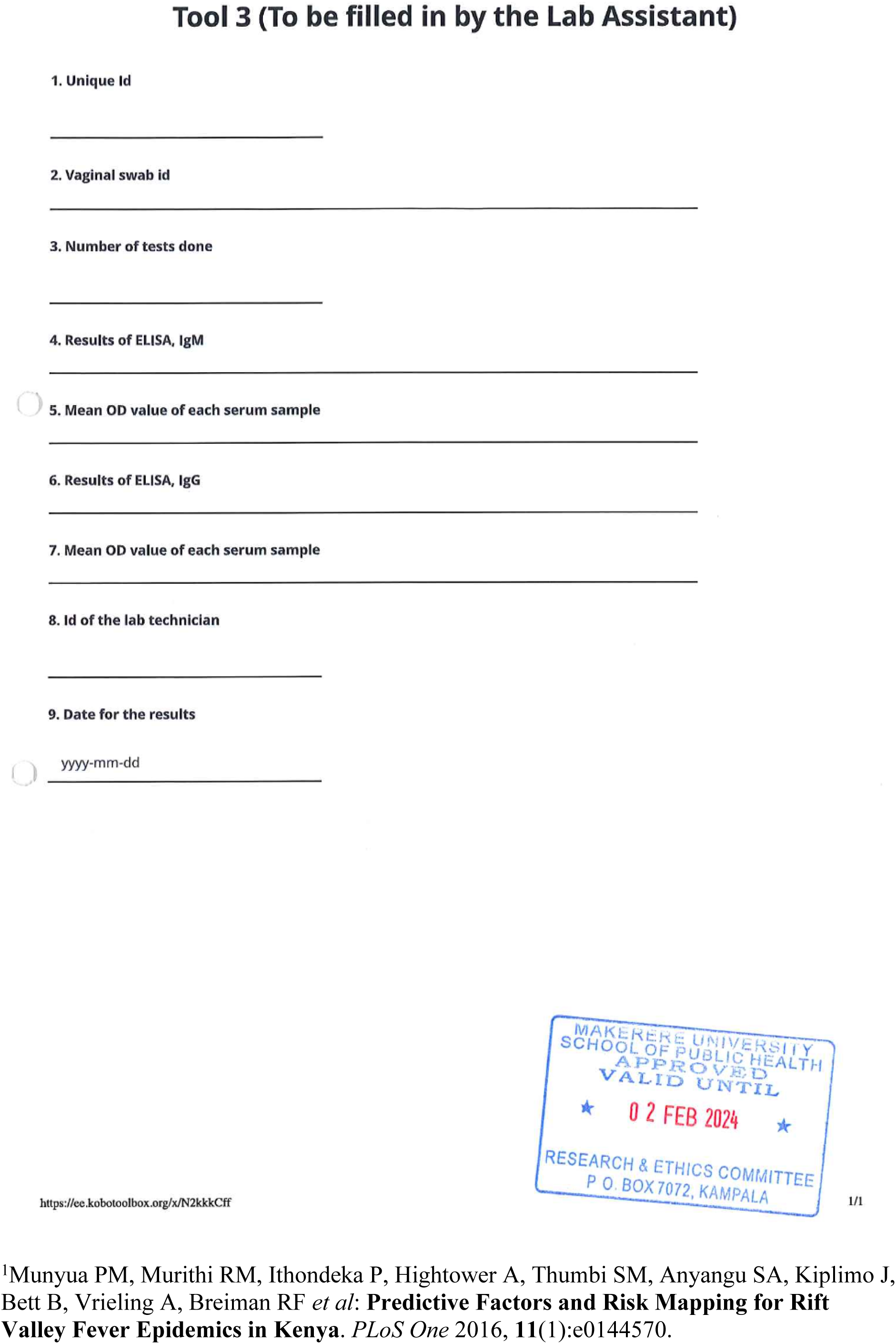
Questionnare (Replicated from Predictive Factors and Risk Mapping for Rift Valley Fever Epidemics in Kenya [1].

**S2 Appendix:**
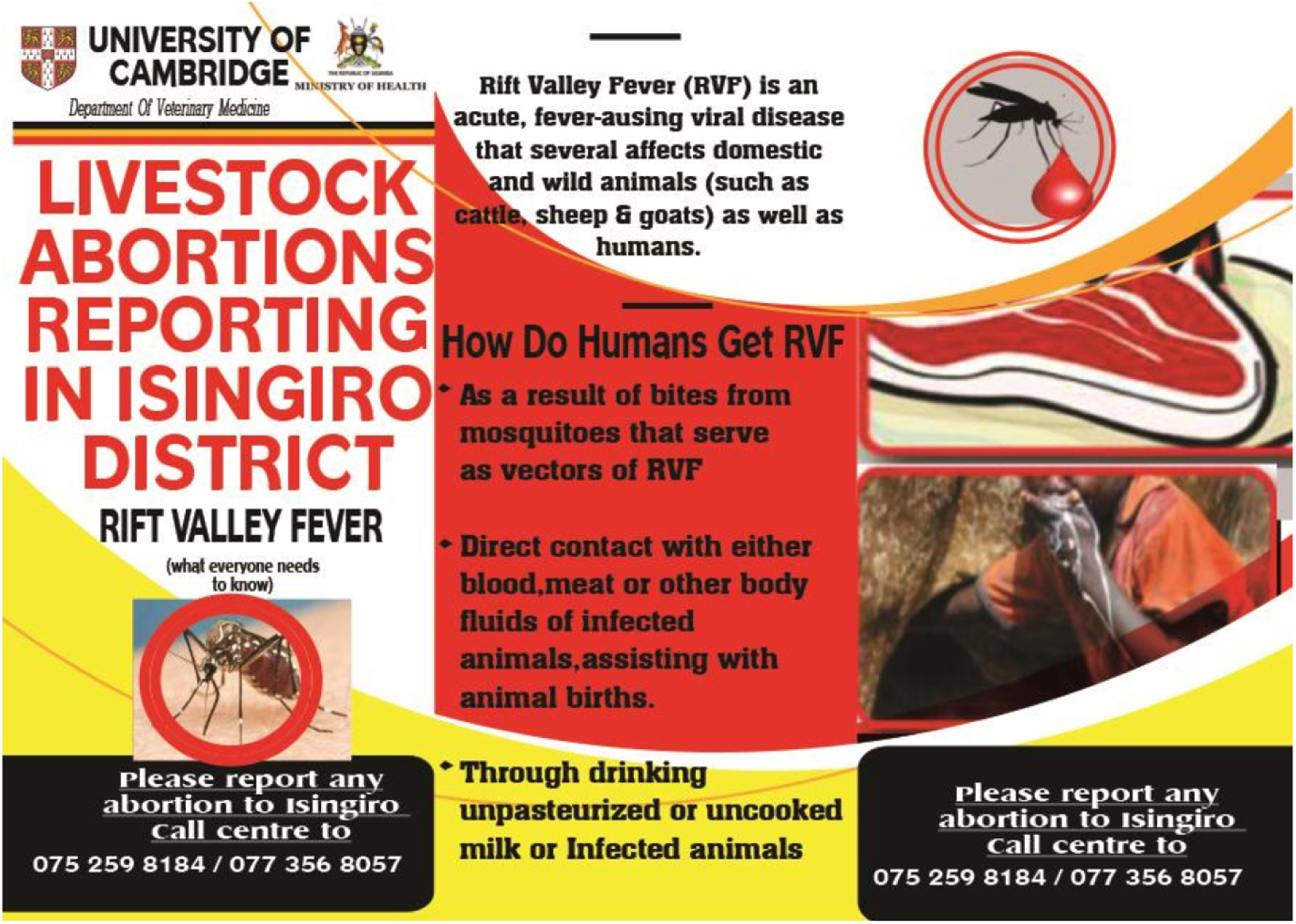
IEC Materials for RVF used during sensitizations (Front page)

**S3 Appendix:**
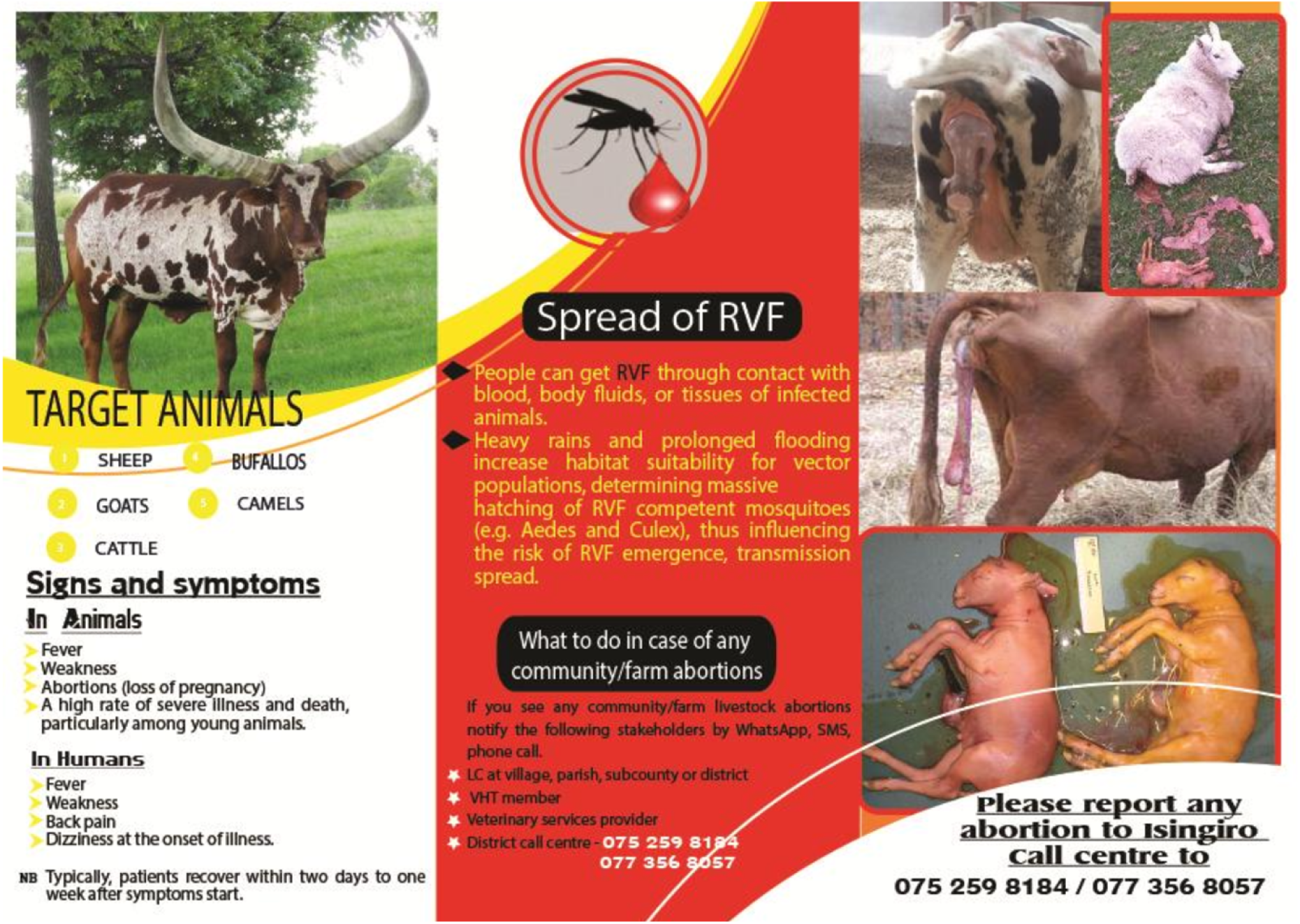
IEC Materials for RVF used during sensitizations (Back page)

## Notes

### Competing Interest Statement

The authors have declared no competing interest.

### Funding Statement

Yes

### Author Declarations

This study is part of the corresponding author's PhD project. We sought and received ethical approval to conduct the research from Makerere University School of Public Health (Reference No: SPH-2022-364), Uganda National Council of Science and Technology (UNCST) (Reference No: A264ES), and Human Biology Research Ethics Committee, University of Cambridge, United Kingdom (Reference No: HBREC.2023.02). Lastly, we sought administrative clearance from Isingiro district local governments’ administration specifically from the Chief Administrative Officer’s office (dated 20th March 2023) for this study to be conducted. Written informed consent from the research participants (Farmers/livestock owners) was obtained for free and voluntary participation in the study. This was after understanding the purpose and costs of the study. During the blood and vaginal swab collection, we also sought written consent for photographing of the animals for potential detection of RVF and follow-up for actions in future. These photos were only for identification of the animals should the vaginal swabs turn positive for RVF for control strategies by Isingiro district Veterinary department or MAAIF. To enhance understanding, we translated all the data collection tools and informed consent into the local language (Runyankole) which was the most spoken language in the Isingiro district. This will ease the understanding of the entire project for the participants to consent. We further kept the information confidential by keeping identifying information (telephone numbers) under key and lock but also coded. The data was analysed and published in aggregate form to avoid identification of individual participants. The data is currently stored under key and lock for the five years after publication as stipulated by UNCST.

